# Reduction in stroke patients’ referral to the ED in the COVID-19 era: A four-year comparative study

**DOI:** 10.1101/2020.05.30.20118125

**Authors:** Saban Mor, Reznik Anna, Shachar Tal, Sivan-Hoffmann Roten

## Abstract

**Introduction:** Current reports indicate that the increased use of social distancing for preventive COID-19 distribution may have a negative effect on patients who suffering from acute medical conditions.

**Aim:** We examined the effect of social distancing on acute ischemic stroke (AIS) patients’ referral to the emergency department (ED)

**Method:** A retrospective archive study was conducted between January 2017 and April 2020 in a comprehensive stroke center. We compare the number of neurologic consultations, time from symptoms onset to ED arrival, patients diagnosis with AIS, number of patients receiving treatment (tPA, endovascular thrombectomy (EVT) or combine) and in-hospital death.

**Results:** The analysis included a total of 14,626 neurological consultations from the years 2017 to 2020. A significant decrease of 58.6% was noted during the months of January-April of the year 2020 compared to the parallel period of 2017. Percent of final AIS diagnosis for the year of 2020 represent 24.8% of suspected cases, with the highest diagnosis rate demarcated for the year of 2019 with 25.6% of confirmed patients. The most remarkable increase was noted in EVT performance through the examined years (2017, n=21; 2018, n=32; 2019, n=42; 2020, n=47).

**Conclusion:** COVID-19 pandemic resulted in routing constraints on health care system resources that were dedicated for treating COVID-19 patients.

The healthcare system must develop and offer complementary solutions that will enable access to health services even during these difficult times.

## Introduction

Currently, the absence of a COVID-19 vaccine or any definitive medication has led to increased use of non-pharmaceutical interventions (NPIs), aimed at reducing contact rates in the population and thereby transmission of the virus.^1^ Two fundamental strategies are possible: (a) mitigation and (b) suppression.

Each policy has major challenges. The strategies differ as to whether they aim to reduce the reproduction number, to below 1 (suppression) – and thus cause case numbers to decline – or merely to slow spread by reducing reproduction number.^1^ NPIs impact depend on the extent to which people respond to instructions, which varies among countries and even communities, with significant spontaneous changes in population behavior even in the absence of government-mandated interventions.

In Israel, several NPI interventions are currently applied: (a) Symptomatic case isolation in home – symptomatic cases are under home isolation until symptoms resolve; (b) Voluntary home quarantine – all household members remain at home for 14 days following identification of a symptomatic case in the household, or when a member returns home from another country; (c) Social distancing of those over 60 years of age – who are required to remain in their households, separate themselves from family members, and avoid hospital and community medical waiting rooms; (d) Social distancing of entire populations – all households reduce contact outside the household, school, or workplace except those affiliated to an approved government institute; and (e) Closure of schools and universities – closure of all schools and universities while shifting to remote learning programs.

Social distancing guidelines were implemented from initial pandemic declaration during the month of December and gradually exacerbated as the pandemic spread and reached a massive global impact by February through April.

These social distancing guidelines indirectly create two isolated populations at high-risk: the chronically ill and voluntary isolated persons who had contact with a verified patient or person returning from abroad. These populations are at increased risk mainly due to difficulty in accessing medical care.

Stroke remains a medical emergency even during a pandemic and although ischemic stroke has been recognized as a complication of COVID-19 the mechanisms and phenotype are not yet understood.^2^

Optimizing outcomes matter even more in these times as patients affected with severe stroke require hospitalization and may potentially be at greater risk of in-patient morbidity and mortality.^3^ Current pandemic compel the elaboration of a more articulate pathway for acute stroke patients, re-routing patient care and allocation of designated pathway for patients with large vessel occlusion.^4^

NPI’s impact on stroke load has yet to be apparent. However, global concerns rises stating that the management of stroke must not be neglected.^5^

## Method

### Study Design and Setting

A retrospective archive study was conducted between January 2017 and April 2020 in a comprehensive stroke center. The hospital serves as a comprehensive stroke center from throughout the north of Israel, and is a referral center for 12 district hospitals, in addition to the Israel Defense Forces Northern Command, the United States Sixth Fleet, United Nations peacekeeping forces and foreign embankment ships stations in the region. The ED contains 105 beds, and about 136,819 patients > 18 years old are treated per year on average. Of these, approximately 5000 patients with suggestive symptoms of AIS are assessed by a neurologist in the hospital annually.

A comparison was made for the months of January-April for the years 2017 to 2020. The study was approved by the institutional review boards of the hospital (#3023–18-RMB).

### Data collection

For each year, data collection was conducted in four steps (Figure 1). First, in order to examine the number of patients admitted with symptoms suggestive of AIS we extracted all neurological consultations from the electronic medical record (EMR). Neurologic consultation can be requested either by the triage nurse or by an ED physician. AIS suspected was defined according to the following key-words: confusion, loss of consciousness, vision abnormalities, dysarthria, facial paralysis, loss of balance, lack of coordination, numbness, weakness, hemiparesis, sensory abnormalities, headaches with abnormalities of gait and diplopia. Neurological consultations of Traumatic head injury, Drug use, Alcohol intoxication, Epileptic seizures, Meningitis, Post herpetic neuralgia, Trigeminal neuralgia, Multiple sclerosis, Parkinson’s disease, Psychosis, Malignancy and Migraine as initial diagnosis were excluded. Second, in order to examine which of the patients had a final AIS diagnosis we collected CT examination results of patients referred from the ED and reviewed the medical records of all patients discharged between the years of 2017 to 2019 diagnosed with AIS except cases of in-hospital stroke. AIS was determined according to the International Classification of Diseases (ICD) 9 codes: 433.01, 433.10, 433.11, 433.21, 433.31, 433.81, and 433.91 (occlusion and stenosis of pre-cerebral arteries), 434.00, 434.01, 434.11, and 434.91 (occlusion of cerebral arteries), 436 (acute but ill-defined cerebrovascular disease). We excluded the following codes 430,431,432 demonstrating hemorrhagic stroke. Third, composing a final patient list by cross linking of records was performed by the first and third author, matching patient’s identification number, case number and exclusion of duplicate records.

**Figure 1.**
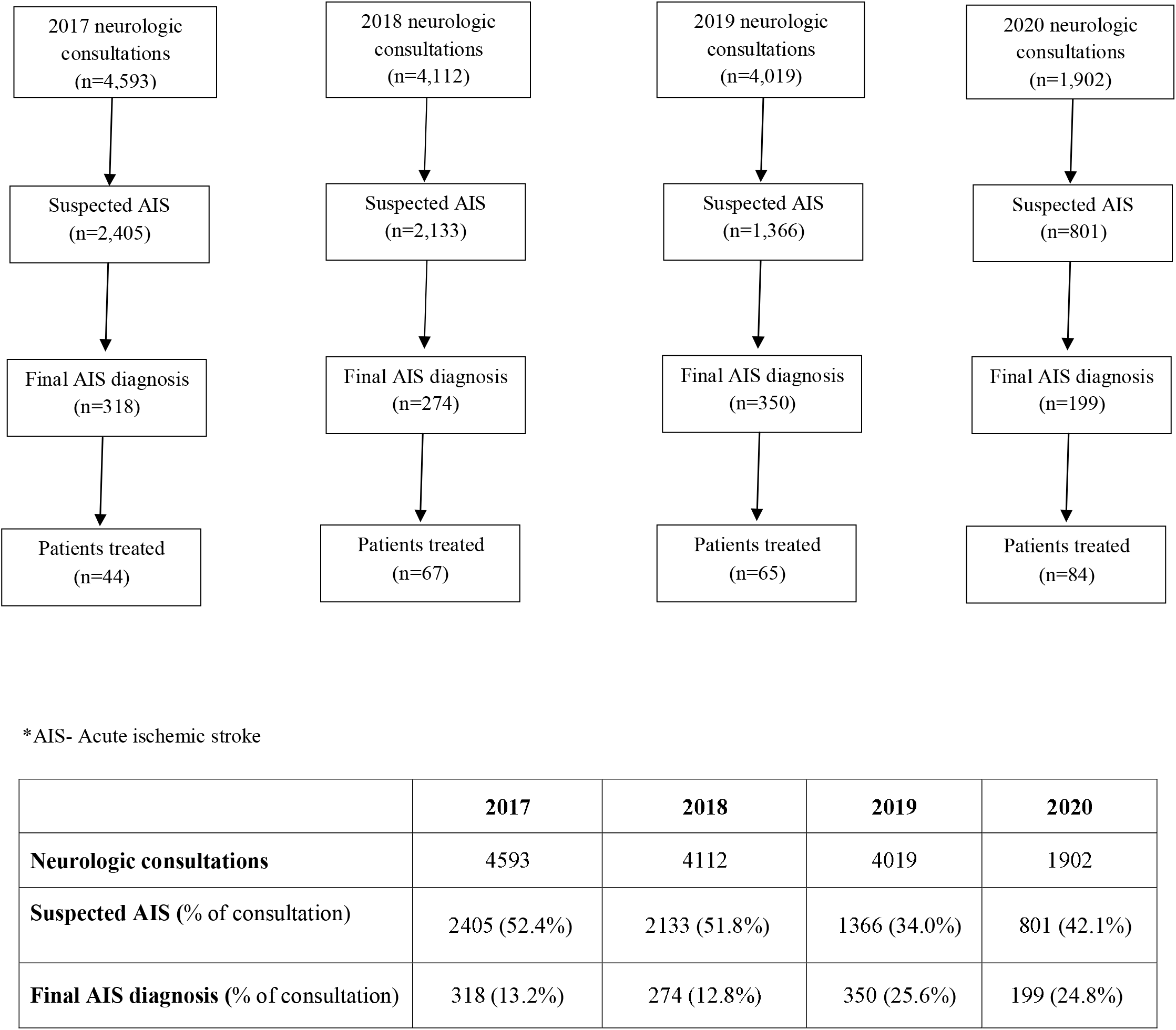
Flow chart

### Statistical Analysis

Continuous variables are reported as means and standard deviations (SD) or medians and interquartile ranges (IQR), as appropriate. Dichotomous variables are reported as proportions. The data analysis was performed using SAS® software (SAS Enterprise Guide 7.1) and Python 3.6.5 software version 2019a.

## Results

The analysis included a total of 14,626 neurological consultations from the years 2017 to 2020, representing 31.4%, 28.1%, 27.5% and 13.0% respectively. A significant decrease of 58.6% was noted during the months of January-April of the year 2020 compared to the parallel period of 2017. Percent of final AIS diagnosis for the year of 2020 represent 24.8% of suspected cases, with the highest diagnosis rate demarcated for the year of 2019 with 25.6% of confirmed patients. (Figure 1) Figure 2 depicts treatment distribution of AIS patients. We found that the number of patients receiving treatment, either tPA or EVT, increased during the year of 2020 (n = 84). With the lowest number of treated patients in the year of 2017 (n = 44), and relative uniformity in the years of 2018 and 2019 (n = 67 and n = 65 respectively). The most remarkable increase was noted in EVT performance through the examined years (2017, n = 21; 2018, n = 32; 2019, n = 42; 2020, n = 47) compared to tPA administration, demonstrating relative uniformity (2017, n = 19; 2018, n = 26; 2019, n = 20; 2020, n = 28).

**Figure 2.**
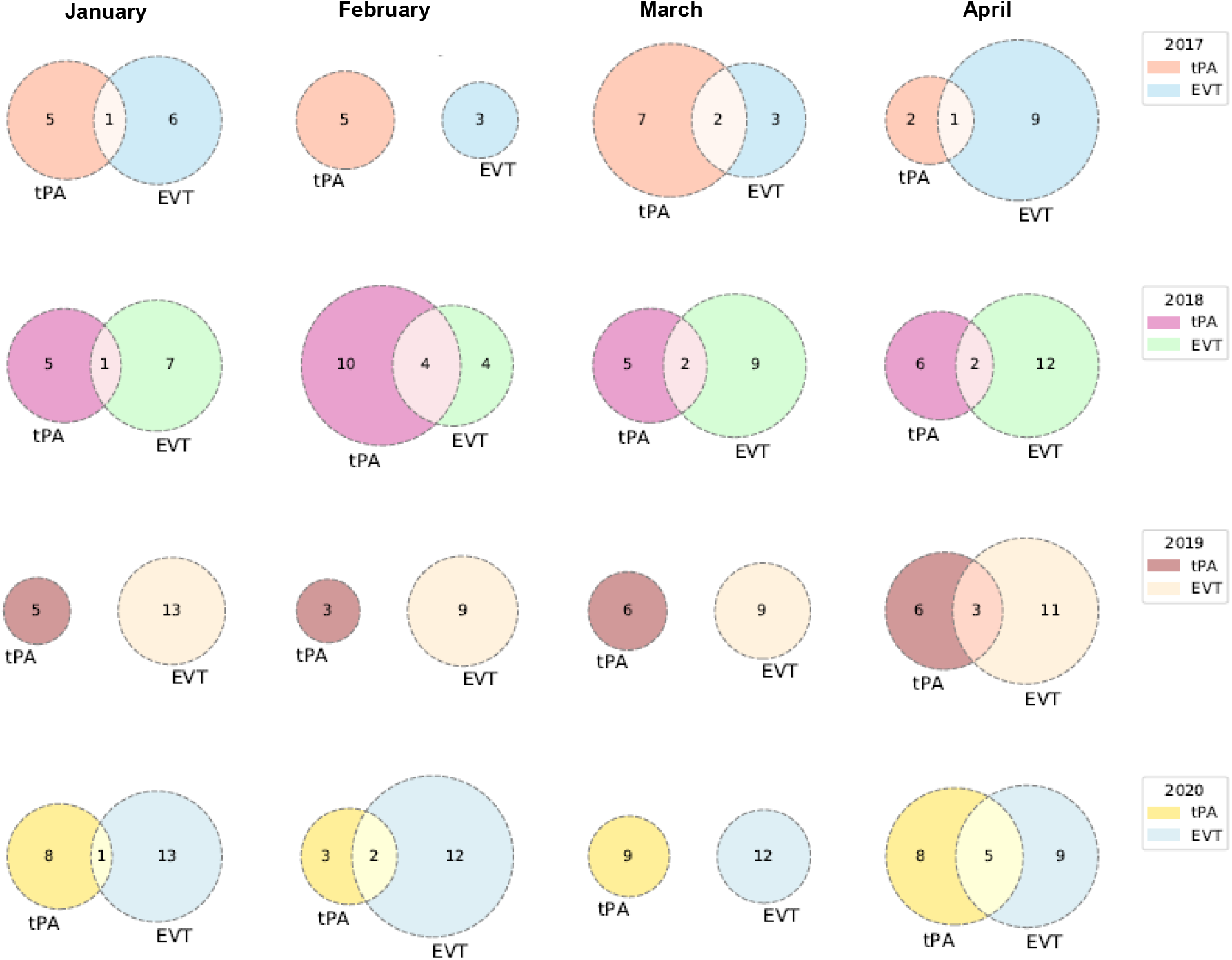
Treatment distribution for AIS patients*. AIS- Acute ischemic stroke; tPA- Tissue plasminogen activator; EVT – Endovascular thrombectomy *Data presented correlates to treatment of AIS patients during the months of January – April for each of the presented years

Patient arrival within six hours from symptom onset was also noted (Figure 3). No significant difference was noticed for the years of 2017 (January, n = 27%; February, n = 27%; March, n = 23% and April, n = 32%), 2018 (January, n = 29%; February, n = 35%; March, n = 42% and April, n = 40%) and 2019 (January, n = 28%; February, n = 34%; March, n = 30% and April, n = 33%). The most remarkable arrival rate was noted for the year 2020 compared to the previous years, with a 100% arrival rate in the month of April.

**Figure 3.**
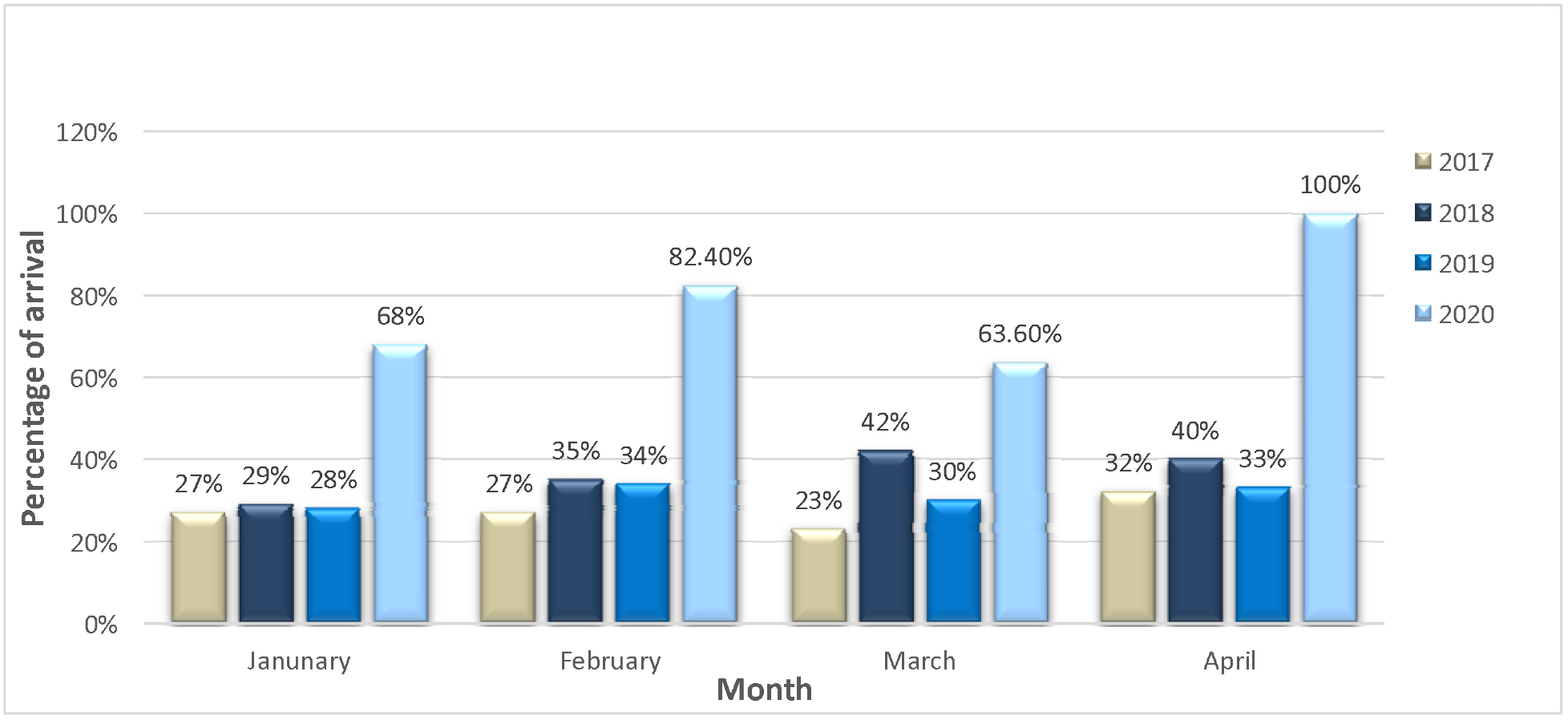
Patient arrival within six hours from symptom onset iva l geo fa r a

For in-hospital death of all-cause mortality, an increase was noted from 2017 (n = 15) to 2018 (n = 32) and 2019 (n = 26), with a subsequential decrease in 2020 (n = 17). Intracranial hemorrhage was found to represent 66.7% of death cases during 2017 and 64.7% in 2020. Lower rates were noted in 2018 and 2019 with 25% and 30.7% respectively (Table 1).

**Table 1.**
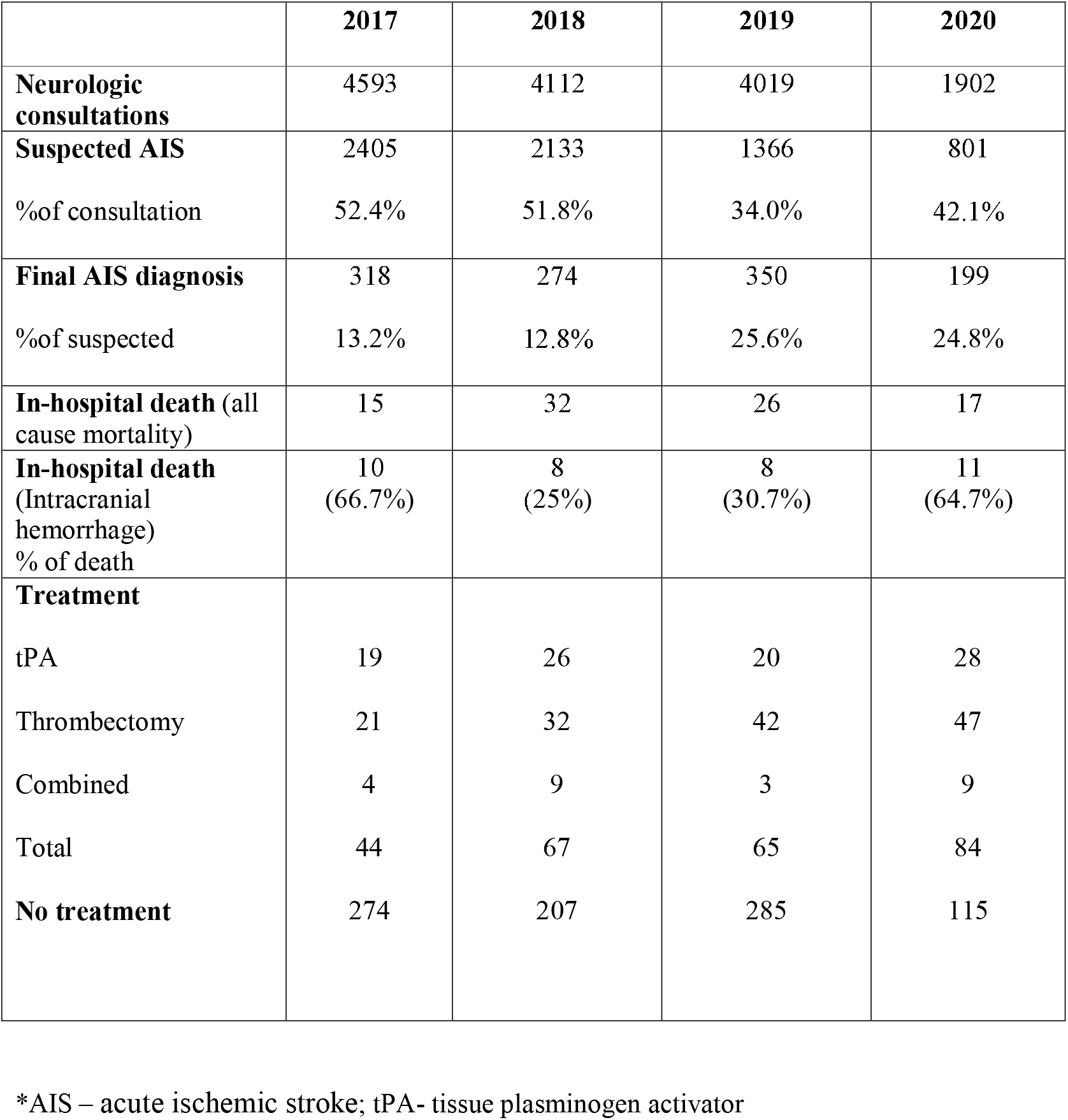
Acute ischemic stroke emergency department workload during January-April.

## Discussion

Currently, the absence of a COVID-19 vaccine or any definitive medication has led to increased use of NPIs, aimed at reducing contact rates in the population and thereby transmission of the virus.^6^ Those NPI’S, primarily based on social distancing has led to a controlled disciplined minimizing viral spread and reduced mortality and morbidity rate.^7^ However, the lack of an anticipated pandemic of such magnitude has forced hospitals to an unprecedented and unexpected channeling of resources into COVID-19 patient treatment. Simultaneously, medical staff and medical services campaigns have urged people to stay in their homes and encourage the use of telemedicine.^8^

Alongside the positive effect of the mitigation strategies undertaken, a number of negative outcomes were also noted.^7^ Among them, a significant reduction in the number of urgent referrals to the ED, one of which is AIS.

It is possible that the fear of contagion at the hospital has discouraged access to emergency medical services, particularly after the media diffused the news that the infection was largely spread across hospitalized patients and healthcare personnel due to the lack of personal protection equipment. Furthermore, second hypothesis is linked to the fact that the emergency medical system was focused on COVID-19 and most healthcare resources were relocated to manage the pandemic. This might have induced an attitude towards deferral of less urgent cases, at both the patient and the healthcare system levels.^8^

In the current study examining AIS patients patterns in the ED during the period of four months for four consecutive years, with consideration to the COVID-19 pandemic, a significant decrease was noticed in the number of neurologic consultations, ED referral for AIS and of those the number of final AIS diagnosis. Numbers illustrate in a significant manner that patients avoided seeking medical care and ED visit. However, the absolute number of patients undergoing treatment has showed an increasing trend during the year of 2020, during pandemic times. A possible explanation may be speculated for symptom severity for those arriving to the ED. Patients willing to seek medical care represented with severs AIS symptoms and required interventional treatment. Second explanation may be attributed to the lower ED arrival rate affiliating to a non-specific medical cause. Thus, neurology teams were able to increase attentiveness for staff guidance and greater attention was placed for each case leading to a more deliberated decision for treatment necessity.

Arrival time distribution demonstrate a significant increase for 2020 compared to the previous year when 100% of patients arrived within 6 hours. A suitable speculation is of severe symptomatic presentation also serving as a possible explanation for early presentation. Although no significant increase in the in-hospital mortality rate was noted, the rate of in-hospital mortality due to intra cranial hemorrhage was significantly higher than parallel periods in the years 2018 and 2019. As such, it may indicate a severe infarct leading to ICH for the elderly arriving with AIS. Craniotomy is usually performed for young patients, especially if the case is a right infarct. In patients with poor prognosis, surgical procedure is usually avoided.^9^

In the present case, the patients arriving may have presented with a massive hemorrhage forming a significant edema, pressing the brain stem. In addition, the Intensive care unit teams preoccupied with the treatment of COVID-19 patients may not had the opportunity to perform a complex procedure such as craniotomy. In contrary to a previous correspondence letter^10^ demonstrating an increase in the number of young patients, COVID-19 positive, presenting with large vessel occlusion, in the current study such increase was not noticed.

## Limitations

This study has several limitations. First, the present study is a retrospective study of medical cases, thus we rely on accurate recordkeeping of others. Secondly, data is available solely for patients arriving to the ED and not for those not referred or their outcomes. Thirdly, there is no long-term follow-up of mortality within 30 days, 90 days and a year as is common in AIS cases. Finally, we cannot completely exclude that a true reduction in the incidence of acute cardiovascular disease as the potential result of low physical stress and widespread prevalence of the resting state during the quarantine, especially in the initial phase of the social containment, might have partly contributed to the lower number of admissions for acute stroke.

## Conclusion

COVID-19 pandemic forced hospitals to an unprecedented and unexpected channeling of resources. Alongside the positive effect of the mitigation strategies taken, a negative effect may be present for patients suffering from acute medical conditions. Paradoxically, lower referral rate and final AIS diagnosis rate were noted parallel to increased treatment percentage. As such, implications may indicate symptom severity and hesitation from seeking medical care. Complementary solutions must be develop by the healthcare system to enable access to health services even during these difficult times.

## Data Availability

The authors have no permission to share the data

